# Use of 1-MNA to Improve Exercise Tolerance and Fatigue in Patients After COVID-19

**DOI:** 10.1101/2021.07.14.21259081

**Authors:** Michał Chudzik, Joanna Kapusta, Monika Burzyńska

**Affiliations:** Medical Center, Saint Family Hospital, Lodz, Poland; Department of Internal Medicine and Cardiac Rehabilitation, Medical University of Lodz, Poland; Department of Epidemiology and Biostatistics, the Chair of Social and Preventive Medicine of the Medical University of Lodz, Poland

**Keywords:** COVID-19, MNA, chronic fatigue syndrome, post-COVID syndrome

## Abstract

**Background:** Coronavirus disease 2019 (COVID-19) is a serious respiratory disease that results from infection with a newly discovered coronavirus (SARS-COV-2). Unfortunately, COVID-19 is not only a short-term infection but that patients (pts) recovering from SARS-CoV2 infection complain of persisting symptoms including: fatigue, diffuse myalgia and weakness, which may lead to chronic fatigue syndrome. There is currently no evidence that nutritional supplements and/or physical exercise can assist in the recovery of pts with chronic fatigue syndrome. 1-Methylnicotinamide (1-MNA) is an endogenic substance that is produced in the liver when nicotinic acid is metabolized. 1-MNA demonstrates anti-inflammatory and anti-thrombotic properties. Therefore, we investigated whether 1-MNA supplements could improve exercise tolerance and decrease fatigue among patients recovering from SARS-CoV-2.

**Methods:** The study population was composed of pts after COVID-19, expressing subjective feelings of limited tolerance to exercise. The selected pts were randomized into two groups: GrM0 – without supplementation; GrM1 – with 1-MNA supplementation. At the beginning of the study (Phase 0), in both groups, a 6-minute walk test (6MWT) was carried out and fatigue assessment with Fatigue Severity Scale (FSS) was performed. After 1 month (Phase 1), a follow up FSS and 6MWT once more were performed in both groups.

**Results:** A significant improvement in the mean distance covered in the 6MWT was noted among the pts in GrM1, compared to those in GrM0. We also noted that in GrM1 the 6MWT distance was significantly higher after 1 month of supplementation with 1-MNA, compared to the beginning of the study (515.18 m in Phase 0 vs 557.8m in Phase 1; p = 0.000034). In GrM1, significantly more pts improved their distance in the 6MWT (23 out of 25 pts, equal to 92%), by a mean of 47 meters, compared to GrM0 (15 of 25 pts, equal to 60%) (p = 0.0061). After one month, significantly more patients in the group without 1-MNA had severe fatigue (FSS ≥ 4) compared to the group with supplementation (GrM1 = 5 pts (20%) vs GrM0 = 14pts (56%); p = 0.008).

**Conclusions:** 1-MNA supplementation significantly improved physical performance in a 6-minute walk test and reduced the percentage of patients with severe fatigue after COVID-19. The comprehensive action of 1-MNA, including anti-inflammatory and anticoagulant effects, as well as activation of the SIRT1 enzyme, may be beneficial for the recovery of patients with persistent symptoms of fatigue and low tolerance to exercise after COVID-19.

## Background

Coronavirus disease 2019 (COVID-19) is a serious respiratory disease that results from infection with a newly discovered coronavirus (SARS-COV-2). The number of COVID-19 survivors is increasing daily. Unfortunately, COVID-19 is not only a short-term infection. Many studies have revealed that patients (pts) recovering from SARS-CoV2 infection complain of persisting symptoms [1] including fatigue, diffuse myalgia and weakness, which may lead to chronic fatigue syndrome [2,3]. Fatigue and limited exercise tolerance are some of the most common symptoms, reported by between 34% and 69% of COVID-19 patients [4,5,6,7]. Many clinical trials have investigated ways to manage these post-COVID symptoms. Unfortunately, no specific treatment for post-COVID fatigue has yet been introduced into clinical practice [8].

Chronic fatigue syndrome (CFS) is characterized by severe and disabling fatigue without a patho-physiologic explanation [9]. According to A.S. Bansal et al. [10] persistent fatigue lasting more than 6 months may be observed in several viral and bacterial infections. Many studies of post-viral fatigue and CFS focus on immune dysregulation and activation caused by changes in cytokine levels [11,12,13]. These changes can lead to microvascular thrombosis and endothelial dysfunction with impaired microcirculatory function, which may explain some post-COVID-19 related symptoms and complications [14,15]. It is likely that multiple and/or synergistic causal mechanisms underlie post-COVID-19 syndrome.

There is currently no evidence that nutritional supplements and/or physical exercise can assist in the recovery of pts with chronic fatigue syndrome. Nonetheless, given that the potential causes of the main symptoms of post-COVID (i.e. limited exercise tolerance and fatigue) include inflammation, endothelial damage, microcirculatory thrombosis and metabolic disorders leading to insufficient production of energy, we should look for supplements that act comprehensively on all these pathogenetic mechanisms. One such molecule is 1-Methylnicotinamide (1-MNA), which is the methylated amide of nicotinic acid (niacin, vitamin B3). 1-Methylnicotinamide is an endogenic substance that is produced in the liver when nicotinic acid is metabolized. 1-MNA demonstrates anti-inflammatory and anti-thrombotic properties. It can have a positive effect on vascular endothelium and improve skeletal muscle energy metabolism [16,17,18,19,20]. Given its multiactivity, the 1-MNA molecule appears to be a good candidate for treating patients with chronic fatigue after COVID-19. Therefore, we investigated whether 1-MNA supplements could improve exercise tolerance and decrease fatigue among patients recovering from SARS-CoV-2.

## Methods

The study population was composed of pts after COVID-19, expressing subjective feelings of limited tolerance to exercise and above 50% greater fatigue compared to their pre-COVID-19 levels. These symptoms must have continued for at least four weeks since the last symptoms of infection. Patients with cardiological and pulmonological complications that could affect the symptoms of reduced exercise tolerance were excluded. Chronic obstructive pulmonary disease and/or asthma patients were also excluded from the study. All pts were given medical examinations including lab-tests for morphology, TSH and creatinine. Only pts with no abnormalities in the lab tests were included in the study. The selected pts were randomized into two groups:

1. GrM0 – without supplementation;
2. GrM1 – with 1-MNA supplementation. MNA supplements were taken once a day at a dose of 58 mg, in the morning after a meal.

At the beginning of the study (Phase 0), pts from both groups were given the following examinations:

- oxygen saturation assessment performed with a pulse oximeter;
- heart rate monitoring;
- dyspnea assessment according to the Borg scale;
- assessment of fatigue with the Fatigue Severity Scale (FSS) test questionnaire.

The FSS is a self-administered questionnaire for assessing the severity of fatigue in different situations over the past week. Each item is rated on a scale from 1 to 7, where 1 indicates strong disagreement and 7 strong agreement. The final score is the mean value of the 9 items. In accordance with previous studies, we considered values ≥4 as indicating severe fatigue [21,22,23].

In both groups, a 6-minute walk test (6MWT) was carried out according to the protocol described in the American Thoracic Society statement on the 6MWT in 2002 [23]. Patients were instructed to walk the greatest distance possible in 6 min, at a self-determined pace, pausing to rest if needed. Walking distance in meters, heart rate (HR), oxygen saturation and dyspnoea rated from 1 to 10 on the Borg CR-10 dyspnoea scale were assessed before and after the test.

After 1 month (Phase 1), a follow up FSS and 6MWT were performed in both groups. The following parameters were assessed and compared with the results in Phase 0:

- distance and the mean difference in distance in 6MWT;
- number and percentage of pts with improved distance in 6MWT;
- FSS score;
- Number and percentage of patients with FSS ≥ 4.

## Statistical Analysis

The data was coded and entered into Microsoft Office Excel and STATISTICA version 13.1. The most important computational methods in the field of descriptive statistics were used in the statistical analysis, including measures of the distribution of measurable features: the arithmetic mean to calculate the average level of the analyzed statistical feature in the population; the standard deviation for the assessment of the dispersion of measurable features; the median to calculate the median value of the feature for the study group when the distribution of the feature deviated from the normal distribution. The structure indexes interpreted in fractions were also calculated (for <100), to assess the ratio of a part of a given statistical population distinguished by a specific level/variant of a feature to the entire studied population. On the basis of these methods, the relationships between the selected statistical features were calculated and the significance of differences in the selected variables was assessed. The χ2 test of independence was used to test the relationship between nominal variables. The Student’s t-test, the U-Mann-Whitney test, the Wilcoxon pairs test and the difference test were used to compare the two groups in terms of the quantitative variables. The Shapiro-Wilk test was used to assess the normality of the distribution. The research hypotheses were verified on the basis of a significance level p ≤ 0.05.

## Results

In total, 50 pts were included in the study: 16 males, 34 females. The groups were randomized: GrM0 – 25 pts, without supplementation; GrM1 – 25 pts with MNA-1 supplementation. The clinical characteristics of the groups in Phase 0 are presented in Table 1.

**Table 1.** Clinical characteristics of the groups GrM0 and GrM1 at Phase 0.

| Parameters | GrM1 | GrM0 | p |
| --- | --- | --- | --- |
| Age | N = 25 | N = 25 | 0.447 |
| Mean | 47.40 | 49.60 |  |
| Standard deviation | 9.73 | 10.55 |  |
| Median | 49 | 50 |  |
| Sex | N=25 | N=25 | 1.000 |
| Female | 17 (0.68) | 17 (0.68) |  |
| Male | 8 (0.32) | 8 (0.32) |  |
| BMI | N = 25 | N = 25 | 0.834 |
| Mean | 26.88 | 27.17 |  |
| Standard deviation | 4.69 | 4.92 |  |
| Median | 25.97 | 26.42 |  |
| Heart rate at rest in Phase 0 | 25 | 25 | 0.352 |
| Mean | 72.40 | 78.64 |  |
| Standard deviation | 11.74 | 1.32 |  |
| Median | 76 | 79 |  |
| Oxygen saturation at rest in Phase 0 | 25 | 25 | 0.522 |
| Mean | 97.28 | 96.96 |  |
| Standard deviation | 2.03 | 4.71 |  |
| Median | 98 | 98 |  |
| Distance in 6 minute walk test in Phase 0 | 25 | 25 | 0.850 |
| Mean | 515.8 | 519.2 |  |
| Standard deviation | 63 | 69 |  |
| Median | 516 | 504 |  |
| Fatigue Assessment According to Fatigue Severity Score in Phase 0 | 25 | 25 | 0.490 |
| Mean | 4.28 | 4.53 |  |
| Standard deviation | 1.38 | 1.16 |  |
| Median | 4.44 | 4.54 |  |
| Dyspnoea on the Borg Scale at rest in Phase 0 | 25 | 25 | 0.128 |
| 0 | 8 (0.32) | 7 (0.28) |  |
| 1 | 3 (0.12) | 2 (0.08) |  |
| 2 | 9 (0.36) | 3 (0.12) |  |
| 3 | 2 (0.08) | 9 (0.36) |  |
| 4 | 2 (0.08) | 2 (0.08) |  |
| 5 | 1 (0.04) | 2 (0.08) |  |
| P | 25 | 25 | 0.205 |

BMI – Body Mass Index; GrM0 – patients without supplementation; GrM1 – patients with MNA-1 supplementation; p – statistical significance.

In each group, there were 4 pts with comorbidities: in GrM1 4 pts had arterial hypertension, in GrM0 3 had arterial hypertension and one pt had Type 2 Diabetes Mellitus. None of the patients had clinical symptoms of heart failure and echocardiography showed no abnormalities in the structure or function of the pts’ hearts. The pts’ medications were not changed during the supplementation period. All patients had correct ambulatory blood pressure. There were no significant differences between the two groups in terms of the mean age, BMI, oxygen saturations, distance in 6MWT, dyspnea on the Borg scale or fatigue assessment on the FSS scale in Phase 0.

## Results after one month with MNA-1 supplementation

### Six-Minute Walk Test

A significant improvement in the mean distance covered in the 6MWT was noted among the pts in GrM1, compared to those in GrM0. Patients with MNA doubled the distance difference after 1 month (Phase 1) compared to GrM0 (mean GrM0 18.14 m vs 38.86 m in GrM1). The results are shown in Figure 1.

**Figure 1.**
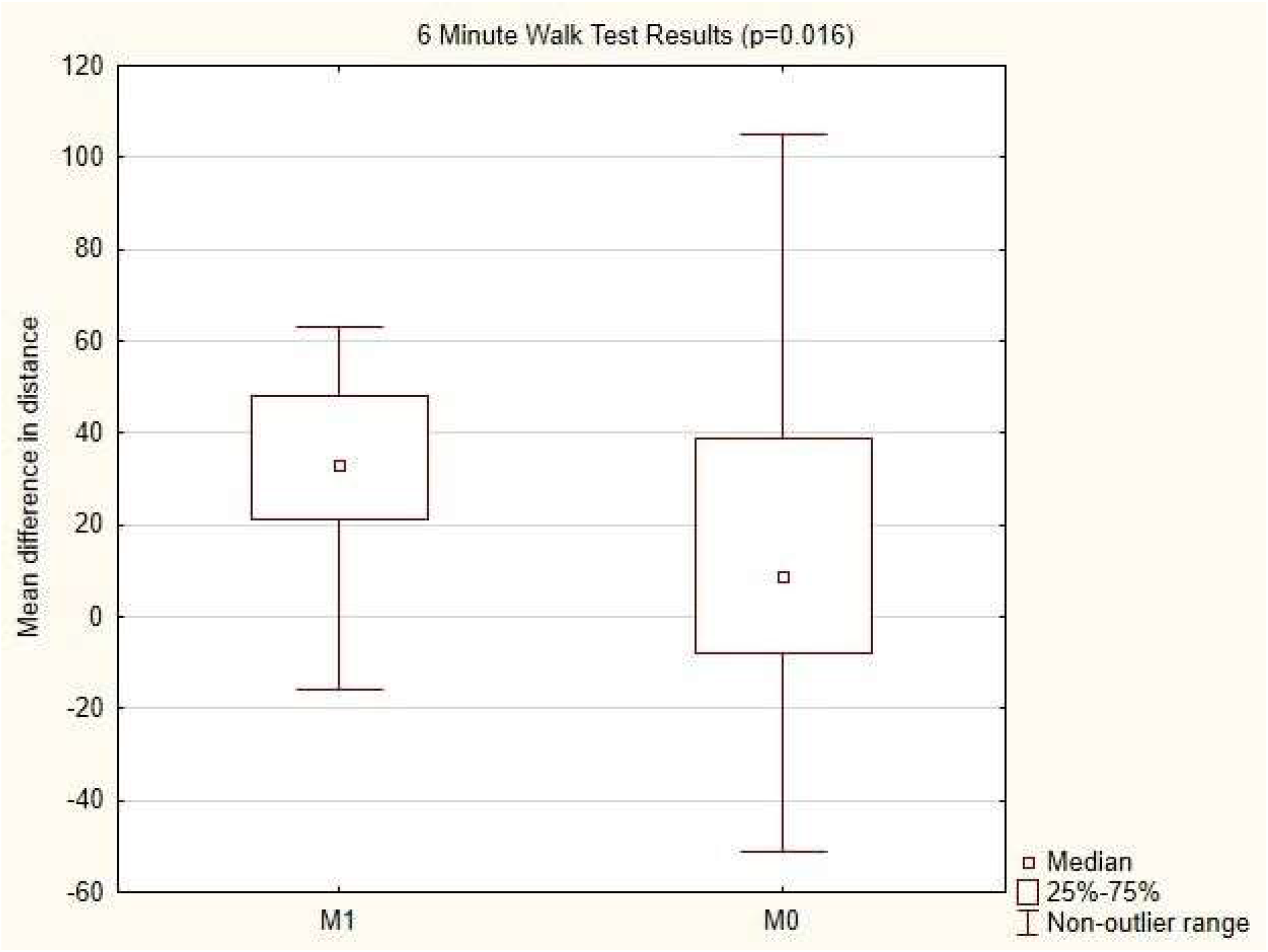
Comparison of mean differences in distance between Group M0 and M1 in 6MWT after 1 month supplementation with 1-MNA. GrM0 – patients without supplementation; GrM1 – patients with 1-MNA supplementation; 6MWT – 6-minute walk test.

We also noted that in GrM1 the 6MWT distance was significantly higher after 1 month of supplementation with 1-MNA, compared to the beginning of the study (515.18 m in Phase 0 vs 557.8m in Phase 1; p = 0.000034). In GrM0 the distance also increased, but the difference was not statistically significant (519.24 m in Phase 0 vs 532.52 m in Phase 1; p = 0.068). The results are shown in Figure 2.

**Figure 2.**
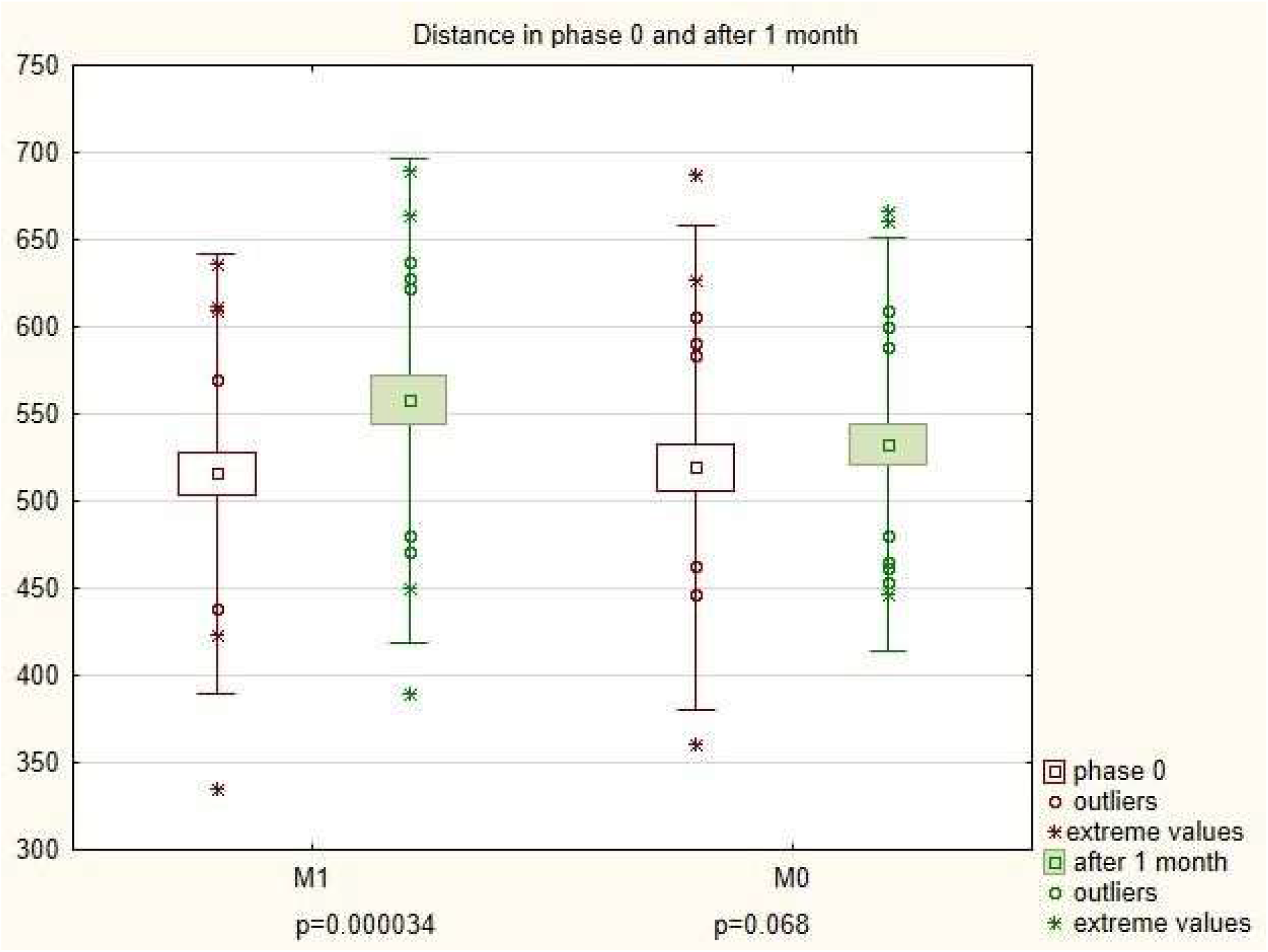
Mean distance in Group M0 and M1 in 6MWT after 1 month of supplementation with 1-MNA.

In GrM1 significantly more pts improved their distance in the 6MWT (23 out of 25 pts, equal to 92%), by a mean of 47 meters, compared to GrM0 (15 of 25 pts, equal to 60%; p = 0.0061). Only 2 pts in GrM1 did not improve their distance. However, these 2 pts reported less fatigue in the FSS.

### Fatigue

Interestingly, after one month both groups reported an increase in fatigue in the FSS (GrM0 FSS = 4.53 in Phase 0 vs FSS = 4.94 in Phase1 and GrM1 FSS = 4.23 in Phase 0 vs FSS = 4.42 in Phase 1). As in the comparison of FSS in Phase 1, the differences between GrM0 and GrM1 were not statistically significant.

In Phase 0, five pts (20%) in GrM1 had an FSS score ≥ 4. In GrM0 the number of patients with an FSS score ≥ 4 was six (24%) (NS). After one month, significantly more patients in the group without MNA had severe fatigue (FSS ≥ 4) compared to the group with supplementation (GrM1 = 5 pts (20%) vs GrM0 = 14pts (56%); p = 0.008).

## Discussion

To the best knowledge of the authors, this is the first study to assess the effect of supplements on the performance parameters of patients after COVID-19. Our results show a significant improvement in walking distance during the 6MWT test following supplementation with 1-MNA. Both the distance and the mean difference significantly increased after 1 month in the group with 1-MNA supplementation. As many as 95% of the pts with 1-MNA supplementation showed an increase in walking distance. Moreover, significantly fewer patients reported significant fatigue, defined as an FSS score ≥4, after one month of supplementation compared to pts without MNA supplementation.

To date, there is still no specific treatment for patients with post-COVID-19 syndrome. The bulk of research work has rightly focused on prevention and treatment of the acute phase of the disease [15]. Only rehabilitation has been considered as a treatment for improving respiratory function, exercise tolerance, fatigue and quality of life in pts after COVID-19 [25,26]. Fatigue and limited tolerance to exercise are the leading symptoms of Long Covid in hospitalized and not-hospitalized sequelae [1,27,28,29]. Fatigue and limited exercise tolerance following viral infection have a very complex pathology. Effective treatment requires supplementation with a preparation that has comprehensive action: protecting the vascular endothelium, as well as having anti-inflammatory and anti-thrombotic effects [11,14].

Many studies show that 1-MNA possesses anti-thrombotic and anti-inflammatory activity and also increases the lifespan of *Caenorhabditis elegans* through SIRT1-dependent mitohormetic effects [16,17,18]. 1-MNA has been found to increase the production of prostacyclin (PGI2) in endothelial cells in rodent models of thrombosis [16]. 1-MNA also increases the bioavailability of NO in the vascular endothelium and regulates the impaired activity of endothelial nitric oxide synthase [19,30]. The endogenous level of 1-MNA rises during physical exercise, and 1-MNA is regarded as a signalling molecule produced in skeletal muscle coordinating energy metabolism [20]. Fatigue, often caused by acute viral infections such as influenza and COVID-19, may be associated with an insufficient endogenous level of 1-MNA [31]. Przyborowski [32] studied the effects of MNA on exercise capacity and the endothelial response to exercise in diabetic mice. Eight-week old mice were given MNA for 4 weeks; their exercise capacity and endurance running were assessed. The MNA-treated mice showed significantly more resistance to fatigue in endurance exercises than the control (mice without supplementation). Schmeisser [18] also noted the beneficial effect of MNA, demonstrating that this compound increases the mean velocity of nematode movement. It has also been shown that in mice muscle activity induces MNA formation, confirming its role in exercise [33]. In humans, methylation of NA metabolites by NNMT as well as MNA supplementation may become versatile approaches to extend healthspan [34].

In an *in vivo* study, Chlopicki et al. [16] found that pharmacological doses of MNA acutely increase secretion of prostacyclin (PGI2) from endothelial cells and may regulate thrombotic as well as inflammatory processes in rodent models of thrombosis. Another positive effect of MNA for endothelium is to enhance the release of nitric oxide from endothelial cells [19]. Ström suggests that MNA could have a beneficial effect of on muscle function, by enhancing the utilization of energy stores in response to low muscle energy availability [20]. Others have proposed that activating SIRT1 has a positive effect on muscle functions. The literature thus provides very significant evidence that MNA acts in a complex way, which could make it a valuable clinical adjunct to the treatment of fatigue and exercise limitation after COVID-19. Given the available evidence, the use of MNA to prevent inflammation damage associated with COVID-19 seems rational.

The 6MWT has long been used in cardiac and pulmonary rehabilitation as an objective assessment of the physical capacity of patients and the effects of treatment, including training programs [35,36]. The test has also been used to study pts after contracting COVID-19 [37.38]. Numerous studies have shown improvements in the 6MWT walking distance among post-hospital patients [39.40,41]. However, there have been no published studies assessing the results for 6MWT in pts without hospitalization.

In our study, population of non-hospitalized pts, the mean distance in the 6MWT of more than 500 meters was much higher than the distances reported for hospitalized patients (333 meters in Ahmed M Abodonya AM et al. [42] or 240 meters in Curci et al. [43]). Of course, this discrepancy may be due to the milder course of COVID-19. In Wonga’s [44] study on pts with mild COVID-19, the mean distance (491 meters) was similar to that in our study.

The difference in the average distance we obtained with 1-MNA supplementation was comparable to the results for pulmonary rehabilitation (44 meters) after COVID-19 reported by Abodonya [42]. In a study by Spielmanns et al. [45], a group of patients after COVID-19 were given 30 treatment sessions over 3 weeks of comprehensive pulmonary rehabilitation. The distance difference in the 6MWT was on average 180 (± 101) meters for the rehabilitated group. This difference is many times greater than we obtained during MNA supplementation. Similarly, in a study by Tazato [46], the improvement in distance was much greater than in our MNA group, but the follow-up time was longer (3 months).

As in the above-mentioned studies, almost all the pts in our study in GrM1 improved their performance in the 6MWT.

Fatigue and significant reductions in tolerance to exercise are among the most common persistent symptoms after COVID-19 [47,48,49]. Fatigue is a common consequence of several viral infections, as shown in the case of EBV by White et al.[50]. It is still unclear why chronic fatigue and the other long-term complications persist in some COVID-19 patients. However, most researchers and clinicians agree that long-term COVID-19 symptoms are associated with the ability of coronavirus to trigger a massive inflammatory response [51].

The FSS is an established research tool [21]. The average score for FSS among the patients in our study was much higher than in the healthy population and comparable with pts with multiple sclerosis (MS) or after a stroke. Fatigue after COVID-19 is thus a significant clinical problem. MNA supplementation did not result in a reduction in the number of people reporting fatigue. However, the low percentage of MNA-treated pts with significant fatigue defined as FSS ≥ 4 was comparable to that in the healthy population (18% in [21] and 20% in our study). This is a very promising result after such a short period of time. There were statistically significant fewer patients with severe fatigue in the 1-MNA group. On the other hand, the percentage of pts in the control group (without MNA supplementation) with severe fatigue was similar to that among pts with MS or after a stroke. The anti-inflammatory effect of 1MNA, which appears to have an impact on fatigue after SARS-Cov-2, may have had a decisive influence on this result.

Currently, there is no evidence that nutrition or physical exercise can contribute to the recovery of patients with fatigue [15]. A clinical trial is ongoing into the use of supplemental creatine to treat Chronic Fatigue Syndrome, but no results have been published so far [8]. The main limitation of the present study was the short observation time. It would be interesting to see if this initial effect would be sustained over a longer follow-up period. It cannot be ruled out that the MNA Group experienced a placebo effect. A randomized MNA vs placebo study on a larger group of patients over a longer observation period is therefore also needed. However, our preliminary results suggest that supplementation with 1-MLA is a promising treatment for Long-COVID in patients without heart or lung disease and without cardiopulmonary complications.

## Conclusions

1. 1-MNA supplementation significantly improved physical performance in a 6-minute walk test and reduced the percentage of patients with severe fatigue after COVID-19.
2. The comprehensive action of 1-MNA, including anti-inflammatory and anticoagulant effects, as well as activation of the SIRT1 enzyme, may be beneficial for the recovery of patients with persistent symptoms of fatigue and low tolerance to exercise after COVID-19.

## Data Availability

No link

